# ENHANCING VENTRICULAR TACHYCARDIA ABLATION OUTCOMES: THE IMPACT OF FUNCTIONAL MAPPING IN CHAGAS CARDIOMYOPATHY

**DOI:** 10.1101/2024.02.08.24302420

**Authors:** Bruno Wilnes, Beatriz Castello-Branco, Gustavo de Araújo Silva, Marina Pereira Mayrink, José Luiz Padilha da Silva, Marco Paulo Tomaz Barbosa, Anna Terra França, Marcos Roberto Queiroz França, Crizianne Rodrigues Santos de Araújo, Reynaldo Castro de Miranda, Antonio Luiz Pinho Ribeiro, Maria do Carmo Pereira Nunes, André Assis Lopes do Carmo

## Abstract

Background: Chagas cardiomyopathy presents a significant public health challenge in Latin America, marked by high rates of ventricular tachycardia (VT) and implantable cardioverter-defibrillator (ICD) interventions. This study explores the efficacy of functional mapping strategies in VT ablation for Chagas cardiomyopathy compared to conventional voltage map-based approaches. Methods: This observational study, involving 66 patients, employed electroanatomic mapping for both epicardial and endocardial regions. In voltage-based mapping (group 1), all patients underwent bipolar substrate mapping with standard scar settings for epicardial and endocardial regions, while functional mapping (group 2) was obtained using a single ventricular extra-stimulus with Late Potential and Isochronal Late Activation Maps associated with annotation of decremental and blocked electrograms. The primary endpoint was post-ablation VT recurrence over a 30-month follow-up. Results: Voltage-based and functional mapping groups presented 42 and 24 patients, respectively. When compared to the voltage-based mapping strategy, functional mapping-assisted procedures were associated with lower post-ablation VT recurrence on Kaplan-Meier’s log-rank test (P=0.045). After multivariate analysis, functional mapping independently associated with a 90.3% reduction in the risk of 30-month VT recurrence (HR: 0.097, 95% CI: 0.012-0.760, P=0.026) compared to voltage-based mapping. Conclusion: Functional mapping in Chagas cardiomyopathy VT ablation is associated with significantly reduced recurrence rates, emphasizing its potential as an effective strategy in this challenging condition. This study provides valuable insights for improving VT ablation outcomes. However, there is a dire need for a large-scale randomized controlled trial to generate high-quality evidence in this field.

**What Is Known:** 1) Functional mapping holds the potential advantage of identifying substrate situated within normal voltage areas, but there is no direct comparison between functional mapping and conventional voltage map-based strategy.

2) Chagas cardiomyopathy, a highly arrhythmogenic disease, is linked to higher rates of post-ablation ventricular tachycardia (VT) recurrences and poor overall prognosis.

**What The Study Adds:** 1) After adjusting for confounding predictors, a functional map-based strategy showed a 90.3% reduction in post-ablation VT recurrence over a 30-month follow-up period.

2) In 26.8% of patients with Chagas cardiomyopathy referred for VT ablation, a periaortic substrate was identified—a location not previously documented in this condition; poor mapping of this region may contribute to ablation failure.

**Graphical Abstract:** 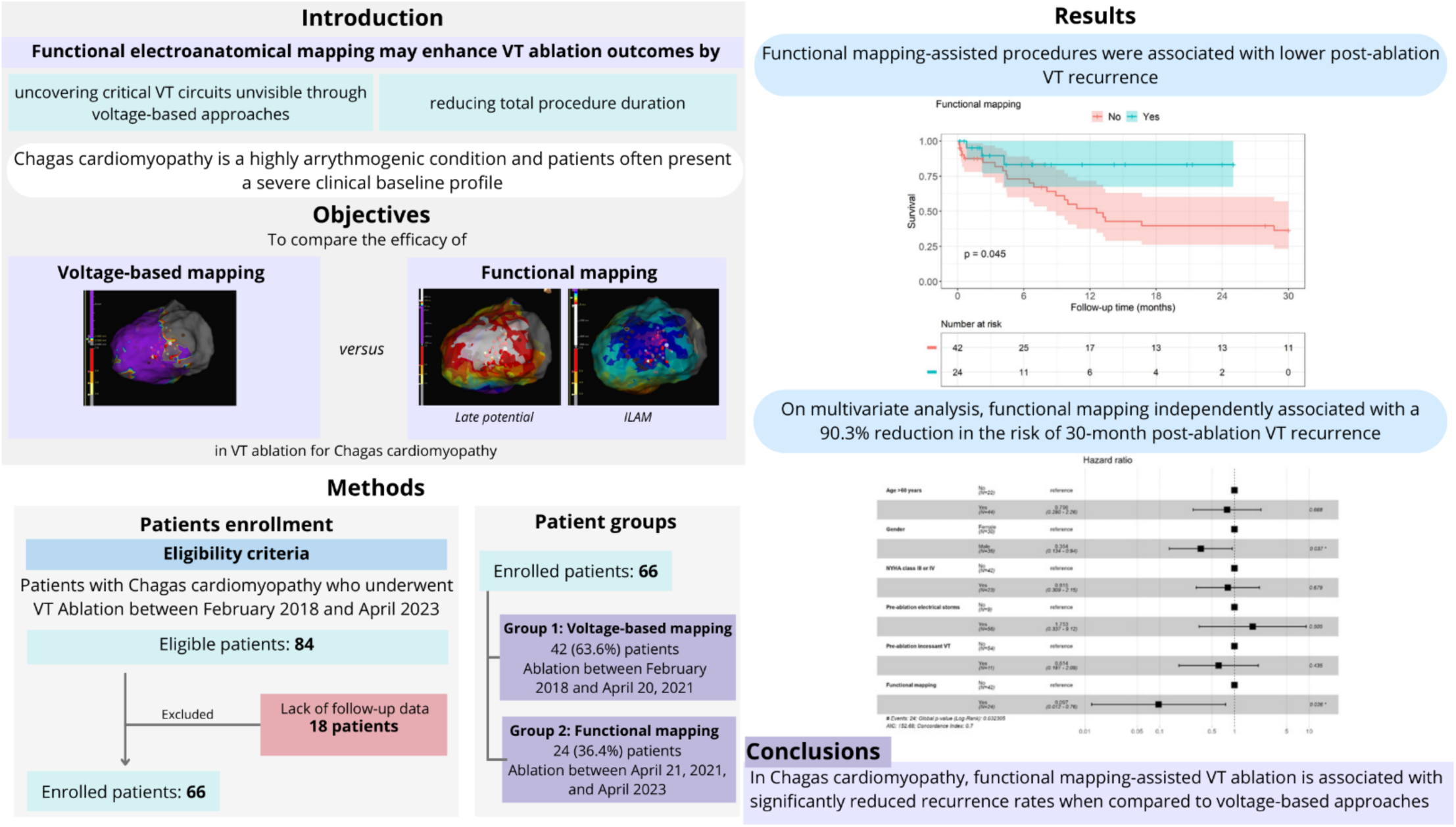

## Background

Chagas cardiomyopathy continues to pose a significant public health challenge in Brazil and Latin America, with an estimated prevalence of 912 cases per 100,000 individuals^1^. The high incidence of ventricular tachycardia (VT) in this population contributes to elevated rates of appropriate implantable cardioverter-defibrillator (ICD) interventions compared to patients with other forms of cardiomyopathy^2–4^. Consequently, there is an urgent need for strategies to enhance VT ablation outcomes in this highly arrhythmogenic cardiomyopathy.

In addition, successful VT ablation involves a series of critical steps. Recent advancements in the field have introduced innovative strategies that focus on the functional properties of the ventricular substrate, offering promising results^5–11^. These strategies hold the potential advantage of precisely targeting substrates situated within normal voltage regions. Consequently, they can lead to a more extensive modification of the substrate area.

The SURVIVE-VT (Substrate Ablation versus Antiarrhythmic Drug Therapy for Symptomatic Ventricular Tachycardia) trial^11^ demonstrated reduced rates of ICD shocks, cardiovascular mortality, and hospitalizations for worsening heart failure through traditional substrate modification. However, there is currently a lack of direct comparisons between functional mapping and the conventional voltage map-based strategy in terms of VT recurrence following catheter ablation. Furthermore, owing to the existing gaps in knowledge surrounding the ideal VT mapping strategy, current guidelines refrain from offering robust recommendations in this context. The objective of this study is to assess the potential association between functional mapping strategies and a reduction in VT recurrence rates following catheter ablation in patients with Chagas cardiomyopathy.

## Methods

### Study design

This is a single-center observational retrospective cohort study, which was performed based on the dataset of a Brazilian reference center specialized in VT management. The report of this study is in line with the STrengthening the Reporting of OBservational studies in Epidemiology (STROBE) recommendations^12^.

### Setting and participants

Patients submitted to VT ablation between February 2018 and April 2023 in our tertiary center were eligible to comprise the analysis. The referred time marks are, respectively, the creation of the dataset and the beginning of the data analysis. Further inclusion criteria were as follows: 1) ventricular tachycardia caused by Chagas cardiomyopathy (CC-VT); 2) clinical follow-up for post-ablation VT recurrence.

Patients were grouped into 2 categories according to the type of electroanatomical mapping performed: 1) voltage-based mapping alone (group 1); 2) functional mapping (group 2). Mapping protocols are comprehensively described in the following sections. Functional mapping techniques were implemented in our service on April 21, 2021, and, due to growing evidence of such techniques enabling a more complete substrate modification,^5,9,13–15^ since its implementation, all VT ablation procedures performed in our service were functional mapping-assisted. Therefore, patients submitted to VT ablation between February 2018 and April 20, 2021, comprised group 1 (voltage-based substrate mapping), and those submitted to VT ablation between April 21, 2021, and April 2023 were assigned to group 2 (functional mapping). The study was approved by the Research Ethics Committee of Federal University of Minas Gerais under the protocol number 0682020300011, and all enrolled patients provided informed written consent.

### Mapping protocol and ablation

All procedures were performed under general anesthesia. Femoral venous access was established, and catheters were positioned in the right ventricle (RV) and the coronary venous sinus. RV programmed electrical stimulation (PES) was carried out to assess the morphology of clinical or targeted VTs, determine inducibility, and evaluate hemodynamic tolerability^16^. The decision to employ intra-aortic balloon pumps (IABP) was based on the PAINESD risk score, and IABP-assisted procedures were recommended when PAINESD ≥15 or at the operator’s discretion^17,18^. Elective IABP placement was preferably scheduled 24 hours prior to the ablation procedure.

Our initial approach primarily involved epicardial access employing a micropuncture needle. Epicardial access was not performed in cases where patients exhibited a high likelihood of significant adhesions. For endocardial access to the left ventricle, we utilized either retrograde arterial and/or transseptal approaches^19^. Endocardial right ventricle (RV) mapping was performed in cases where VT morphology suggested its origin or when epicardial mapping revealed scarring on this chamber. Before endocardial access to the left ventricle, we administered systemic anticoagulation through intravenous heparin, targeting an activation clotting time within the range of 250 to 350 seconds^20^.

Electroanatomic mapping was performed for all patients. For patients undergoing voltage-based substrate mapping (group 1), we employed EnSite Precision (2-2-2 Livewire or Advisor HD Grid Mapping Catheter Sensor, Abbott Park, IL) or CARTO (Thermocool, Biosense Webster, Diamond Bar, CA). EnSite Precision (2-2-2 Livewire or Advisor HD Grid Mapping Catheter Sensor, Abbott Park, IL) was utilized for all patients undergoing functional mapping (group 2) due to the availability of recorded electrograms, enabling the concurrent construction of other functional maps during ablation procedures.

During the mapping process in the epicardial or endocardial left ventricle, RV pacing was the preferred method. Initially, mapping and ablation in VT were deferred, even if the VT was hemodynamically tolerated. Exceptions were made for patients with incessant VT or those who remained inducible after substrate modification. Regions containing His electrograms and phrenic nerve locations were identified and marked with separate color-coded tags.

### Voltage-based substrate mapping

Traditional substrate mapping techniques primarily involve the assessment of ventricular scar substrate within the intrinsic rhythm^21^. In group 1, all patients underwent bipolar voltage mapping with standard scar settings for epicardial and endocardial regions. During bipolar voltage mapping, the criteria for defining normal tissue were >1.5 mV for both left ventricle (LV) and RV endocardium^22^, and >1.0 mV for the epicardial surface^23^, all during intrinsic rhythm. Dense scar was identified as <0.5 mV in the LV endocardium^22^. Additionally, a unipolar voltage (UV) threshold of <8.27 mV was considered for all LV endocardial sites^24^, with the exception of LV septum and periaortic substrates, for which the cut-offs <4.8 mV and <7.5 mV were, respectively, implemented^25,26^. Finally, a RV endocardial unipolar voltage threshold of <3.9 mV defined RV epicardial abnormal substrates^27^. Table 1 summarizes all bipolar and unipolar references employed for voltage map construction. Areas displaying fractionated or late potentials (LP) were marked with color-coded tags. Mapping procedures were deemed complete when the entire region of interest was fully mapped, and all scar borders were clearly delineated.

**Table 1.** Voltage reference used for voltage map construction.

|  | **Left ventricle**<br>**endocardium** | **LV endocardial**<br>**septum** | **Periaortic**<br>**endocardium** | **Right ventricle** | **Epicardial** |
| --- | --- | --- | --- | --- | --- |
| Bipolar | 0.5 – 1.5 mV | 0.5 – 1.5 mV | <1.5 mV | <1.5 mV | <1.0 mV |
| Unipolar | <8.27 mV | <4.8mV | <7.5 mV | <3.9 mV | - |

LV: Left Ventricle; mV: Millivolts.

The ablation procedure was carried out at the optimal ablation sites, which were identified based on the regions featuring fractionated, delayed, or abnormal electrograms as marked during the intrinsic rhythm mapping. Additionally, the entire scar, as defined by the voltage-based map created through 3D mapping, was also targeted.

The primary endpoint in this group was the elimination of all abnormal potentials, which was documented through the absence, or by significant modification, of these potentials as assessed through visual inspection. Following the ablation session, PES was performed to evaluate the inducibility of any VT^16^. The areas with abnormal potentials were remapped after ablation to confirm the extent of modification.

### Functional mapping

In this group, all maps were obtained using a single extra-stimulus delivered from the RV, timed 20 ms above the effective refractory period. Each electrogram was manually annotated with the objective of incorporating the latest local activation time. Areas displaying near-field electrogram decrement or block were distinguished with color-coded tags, irrespective of the voltage recorded.

Activation mapping of late potentials was initially performed. LPs were identified as isolated high-frequency local electrograms occurring after the offset of the terminal portion of the QRS complex and were represented in the last isochron (depicted arbitrarily by the color white). The TurboMap tool (EnSite Precision, Abbott Park, IL) was used to create other maps, including isochronal late activation, obtained from both extra-stimulation and intrinsic rhythm. These maps provided additional information for ablation target selection.

The voltage-based substrate map was generated using the conventional low voltage criteria mentioned earlier, which delineated areas of dense scar in both the endocardial and epicardial regions. Isochronal Late Activation Mapping (ILAM)^28,29^ was configured with a time window commencing at the earliest site of activation and extending to the most delayed site of activation that was mapped. Activation times were displayed with the use of eight evenly spaced activation isochrones. Isochronal crowding was characterized by the localized slowing of propagation, signified by the convergence of multiple isochrones, with more than two isochrones present within a 1 cm^2^ radius.

In the functional map group, the color tags on the late potential areas of the 3D map served as the primary radiofrequency (RF) ablation targets. Concurrently, other maps were constructed using the TurboMap tool (EnSite Precision, Abbott Park, IL). RF applications were performed on areas featuring tagged abnormal local electrograms, characterized by decremental properties or block, and areas with extra-stimulus-induced isochronal crowding identified through ILAM.

Such procedures were performed to achieve epi-endocardial substrate modification, specifically focusing on the elimination of LPs, with the goal of rendering the patient non-inducible for any VT by the conclusion of the procedure. Following the ablation, post-ablation remapping with extra-stimuli was performed to assess the impact of the ablation on the targeted LPs. This remapping served to verify the absence of residual slow conduction and late activation in the treated areas. Additional ablation was carried out if the remapping revealed the persistence of LPs. PES was repeated, using up to the fourth extra-stimuli, to confirm the non-inducibility of any VT.

In cases where VT remained inducible after these efforts, activation mapping was performed during VT. Targeted ablation of the protected isthmus was pursued as dictated by the patient’s hemodynamic tolerance and the specific characteristics of the VT, as shown in Figure 1.

**Figure 1.**
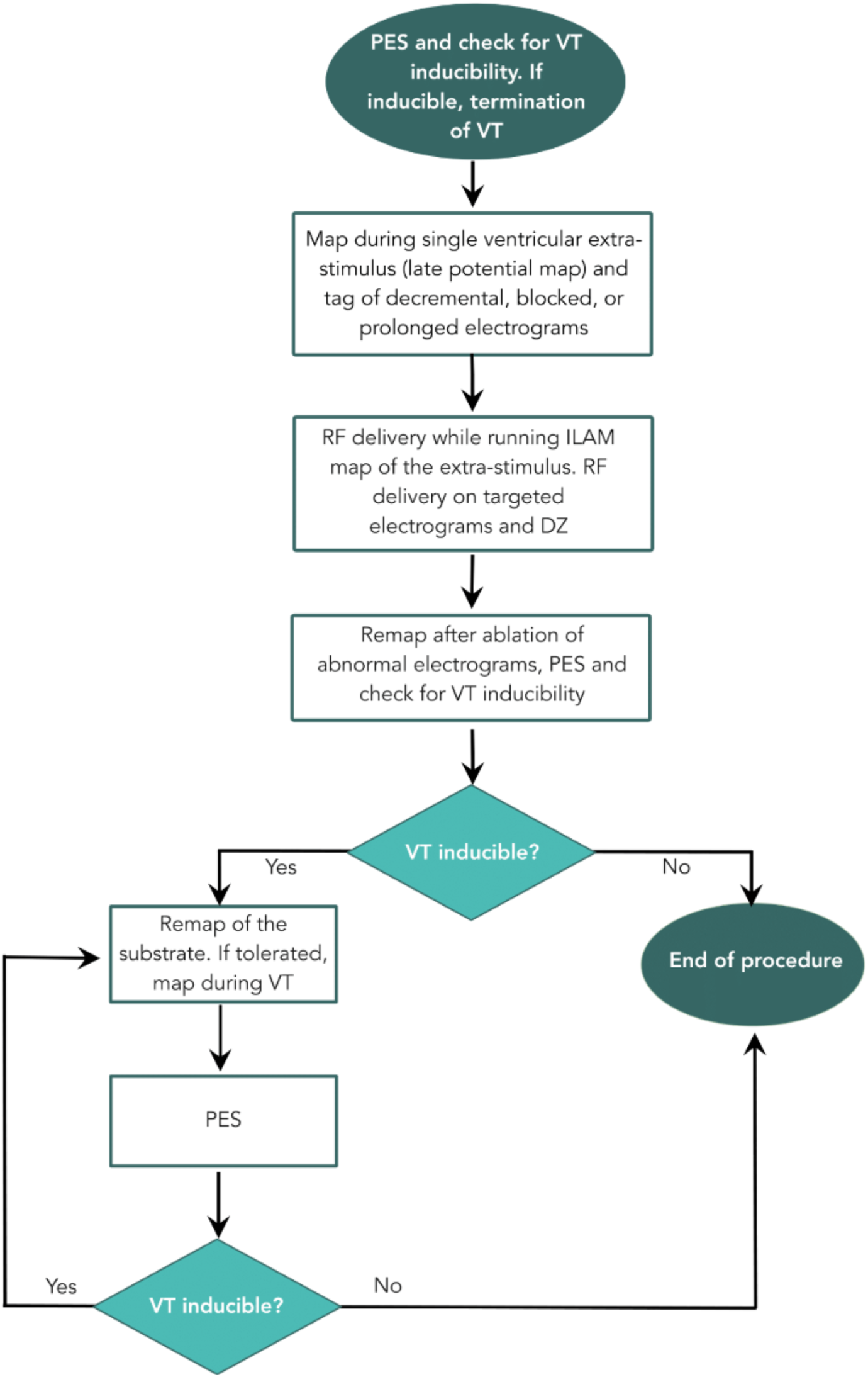
Flowchart illustrating the functional mapping and ablation protocol for ventricular tachycardia ablation performed in group 2. VT: Ventricular tachycardia. RF: Radiofrequency. DZ: Deceleration zones. PES: Programmed electrical stimulation.

At the conclusion of the procedure, intrapericardial corticosteroids were administered to prevent pericarditis^30,31^. We diligently recorded all the complications associated with the ablation procedure.

### Outcome definition

The primary outcome of this study was VT recurrence over a 30-month follow-up period, defined as the first occurrence of sustained VT during post-ablation follow-up. In our study, follow-up ended at 30 months post-procedure, after death, or after the manifestation of the endpoint “VT recurrence”.

### Statistical analysis

Continuous variables were expressed as median and interquartile range (25%-75%) and compared using the Mann-Whitney U test. Categorical variables were presented as absolute numbers and percentages and compared using the chi-square test.

Furthermore, we performed risk estimations using a multivariate Cox proportional-hazards model to identify the effect of functional mapping-assisted ablation in the risk of post-procedure VT recurrence, after adjustment to clinically relevant variables that could perform as confounding parameters. Three predetermined parameters that comprise the PAINESD Score^17^ (age, pre-ablation NYHA functional class, and pre-ablation VT electrical storm) were initially selected to enter the multivariable model according to the PAINESD format (“age >60 years”, “pre-ablation NYHA functional class 3 or 4”, and “pre-ablation VT electrical storm”). We performed Kaplan-Meier curves to graphically analyze each variable included in the multivariate model; for numerical variables, log hazard splines-adjusted curves were also performed (Supplementary Figures S1-S2). Pre-ablation LVEF was analyzed according to PAINESD Score format (“pre-ablation LVEF <25%”), and exhibited significant nonlinearity at the splines curve (Supplementary Figure S1); since it was noticed that this predictor stratified the study population into two subpopulations with very distinctive risk profiles, “pre-ablation LVEF <25%” was included as a strata variable in the final model. As a culminating step in multivariate analysis, we added functional mapping to the model and performed the likelihood ratio test (LRT)^32^ to assess its incremental value in enhancing the model’s overall performance.

Additionally, we conducted tests for proportional hazards, interaction assumptions, and multicollinearity among variables, and found no violations. To better illustrate each parameter’s effect in the multivariate model, we created forest plots containing the variables included in the Cox models. Additionally, a cumulative survival curve for the occurrence of post-ablation VT recurrence was performed by the Kaplan-Meier method and compared using the log-rank test. The cumulative survival curve was accompanied by a corresponding “number at risk” description. A value of P<0.05 was considered statistically significant. All analyses were performed in SPSS version 24 (SPSS Inc., Chicago, Illinois) and in R (R Core Team, 2023).

## Results

### Study population

A total of 84 patients underwent CC-VT ablation between February 2018 and April 2023. However, 18 patients lacked clinical follow-up data and were excluded from the study. The final analysis included 66 patients who underwent CC-VT ablation between February 2018 and April 2023 presenting complete data regarding clinical follow-up for post-ablation VT recurrence (Figure 2). Furthermore, we compared the included and excluded populations to examine group composition for any significant differences, and there were no statistical distinctions. Supplementary Table S1 summarizes these comparisons.

**Figure 2.**
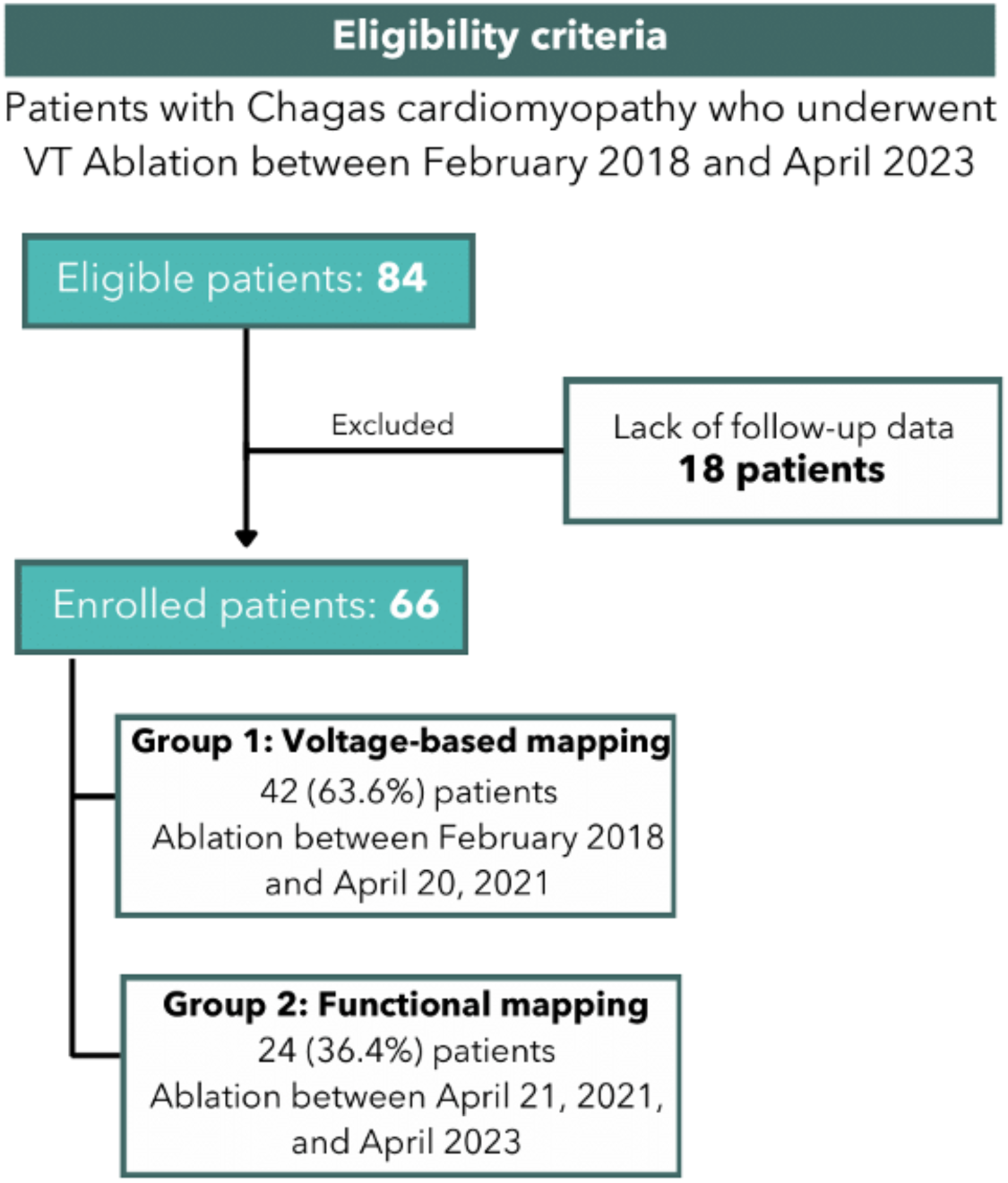
Fluxogram showing the eligibility criteria and patients’ enrollment process. VT: Ventricular tachycardia.

Functional electroanatomic mapping was performed in 24 patients (group 2), constituting 36.4% of the total cohort. Comprehensive baseline clinical characteristics of groups 1 and 2 are detailed in Table 2. The median age was 64.2 (57.1-72.8) years, with 44 (66.7%) patients presenting age >60 years. Additionally, 36 individuals (54.5%) were males, and the median left ventricular ejection fraction (LVEF) before ablation was 37.5% (29.8-47.0). Notably, LVEF was <30% in 16 patients (24.2%), and <25% in 7 (10.6%) individuals.

**Table 2.** Baseline Characteristics According to the Performing or not of Functional Mapping for VT catheter ablation in Chagas Cardiomyopathy.

| Parameters | Group 1 (n = 42) |  | Group 2 (n = 24) |  | P-Value |
| --- | --- | --- | --- | --- | --- |
|  | N | Descriptors | N | Descriptors |  |
| Follow-up time |  |  |  |  |  |
| Follow-up until study end-point, median (P25-P75), months | 42 | 8.5 (1.7-30.0) | 24 | 5.1 (1.9-13.5) | 0.211 |
| Follow-up time until death, 30 months post-ablation, or follow-up loss, median (P25-P75), months<sup>#</sup> | 42 | 10.4 (2.1-30.0) | 24 | 5.1 (2.0-13.5) | 0.085 |
| Age, median (P25-P75), years | 42 | 65.2 (58.5-73.5) | 24 | 63.0 (55.3-69.0) | 0.317 |
| Age >60 years (%) | 42 | 31 (73.8%) | 24 | 13 (54.2%) | 0.103 |
| Male gender (%) | 42 | 20 (47.6%) | 24 | 16 (66.7%) | 0.135 |
| Diabetes mellitus (%) | 14 | 2 (14.3%) | 18 | 1 (5.6%) | 0.568 |
| Chronic obstructive pulmonary disease (%) | 14 | 0 (0%) | 18 | 1 (5.6%) | 1.000 |
| Coronary artery disease (%) | 14 | 1 (7.1%) | 18 | 0 (0%) | 0.438 |
| Implanted device (%) | 41 | 39 (95.1%) | 22 | 21 (95.5%) | 1.000 |
| Pre-ablation NYHA class |  |  |  |  |  |
| NYHA 1 (%) |  | 4 (9.8%) |  | 6 (25.0%) |  |
| NYHA 2 (%) |  | 19 (46.3%) |  | 13 (54.2%) |  |
| NYHA 3 (%) | 42 | 11 (26.8%) | 24 | 3 (12.5%) | 0.191 |
| NYHA 4 (%) |  | 7 (17.1%) |  | 2 (8.3%) |  |
| Pre-ablation LVEF, median (P25-P75), % | 42 | 37.5 (28.8-45.3) | 24 | 38.0 (30.3-47.8) | 0.739 |
| Pre-ablation LVEF <30% (%) | 42 | 11 (26.2%) | 24 | 5 (20.8%) | 0.625 |
| Pre-ablation LVEF <25% (%) | 42 | 4 (9.5%) | 24 | 3 (12.5%) | 0.699 |
| Pre-ablation PAINESD score, median (P25-P75) | 14 | 8 (5-14) | 18<sup>1</sup> | 8 (6-9) | 0.896 |
| Pre-ablation antiarrhythmic drugs |  |  |  |  |  |
| Amiodarone (%) | 42 | 39 (92.9%) | 24 | 22 (91.7%) | 0.896 |
| β-blockers (%) | 42 | 40 (95.2%) | 24 | 24 (100.0%) | 0.066 |
| Propafenone (%) | 42 | 1 (2.4%) | 24 | 0 (0.0%) | 0.176 |
| Pre-ablation incessant VT (%) | 42 | 9 (21.4%) | 23 | 2 (8.7%) | 0.302 |
| Pre-ablation electrical storms (%) | 41 | 38 (92.7%) | 24 | 18 (75.0%) | 0.066 |
| Number of pre-ablation appropriate ICD therapies, median (P25-P75) | 41 | 66 (28-101) | 20 | 42 (15-81) | 0.176 |

Group 1: patients submitted solely to substrate mapping. Group 2: patients in which functional mapping was performed. <sup>#</sup>For this metric, when patients have suffered VT recurrence and death, only the outcome "death" was considered. \*P-Value < 0.05. Sample distribution was analyzed by the Kolmogorov-Smirnov test. Categorical variables were compared using the Chi-square test or Fisher's Exact test when appropriate. Numerical variables were compared using Student's t test or Mann-Whitney U test, according to sample distribution. LVEF: Left Ventricle Ejection Fraction. NYHA: New York Heart Association functional class. VT: Ventricular Tachycardia. LV: Left Ventricle. SD: Standard Deviation. P25-P75: Percentile 25 - Percentile 75. PAINESD Score: chronic obstructive pulmonary disease, age >60 years, coronary artery disease, NYHA class 3 or 4, LVEF <25%, VT storm, and diabetes mellitus.

Assessments regarding pre-ablation data of New York Heart Association (NYHA) functional class, incessant VT, and electrical storms were available for 65 individuals (98.5%). Among these, 23 (35.4%) were classified as NYHA class 3 or 4, and 9 (13.8%) as NYHA class 4. Considering pre-ablation incessant VT, 11 patients (16.9%) exhibited this abnormal rhythm, while the remaining 54 (83.1%) did not. Also, pre-ablation electrical storms were observed in 56 patients (86.2%). Information about post-ablation complications and intra-ablation need for circulatory support was available for 63 patients (95.5% of the study group). Notably, only 5 individuals (7.9%) experienced post-procedural complications, and an equivalent number of patients (7.8%) required circulatory assistance during VT ablation.

### Ablation and topography of scarring tissue

#### Epicardial mapping

Epicardial mapping was performed in 57 (86.4%) patients, revealing that 52 (91.2%) presented epicardial scars. The approximate topography of epicardial scars was available for 28 individuals (42.4%). Among them, all 28 patients (100%) exhibited epicardial scars in the left ventricle (LV), 2 (7.1%) in the right ventricle (RV), and 2 (7.1%) displayed epicardial scars in both LV and RV.

#### Left ventricular endocardial mapping

Left ventricular endocardial mapping was constructed in 56 (83.6%) patients. Of these, 32 (57.1%) individuals showed endocardial scars in LV. Specifically, scars were observed in various regions: 20 (35.7%) individuals had scars in the basal lateral region, 15 (26.8%) in the periaortic area, 13 (23.2%) in the infero-basal region, 6 (10.7%) in the septal region, and 4 (7.1%) in the apical region.

#### Right ventricular endocardial mapping

Endocardial mapping of the RV was performed in 15 (22.7%) individuals, revealing that 6 (40.0%) of them exhibited RV scars. The distribution of RV endocardial scars was as follows: 4 (26.7%) individuals displayed scars in the septum, 3 (20.0%) in the outflow tract, and 1 (6.7%) in the free wall.

Table 3 describes the incidence of each scar topography between groups.

**Table 3.** Ablation Parameters and Post-Ablation Complications According to the Performing or not of Functional Mapping for VT catheter ablation in Chagas Cardiomyopathy.

| Parameters | Group 1 (n = 42) |  | Group 2 (n = 24) |  | P-Value |
| --- | --- | --- | --- | --- | --- |
|  | N | Descriptors | N | Descriptors |  |
| Ablation parameters |  |  |  |  |  |
| Circulatory assistance (%) | 41 | 3 (7.3%) | 22 | 2 (9.1%) | 1.000 |
| Epicardial scars (%) | 36 | 35 (97.2%) | 21 | 17 (81.0%) | 0.056 |
| LV endocardial scars (%) | 33 | 14 (42.4%) | 23 | 18 (78.3%) | 0.008* |
| LV endocardial periaortic scar (%) | 33 | 4 (12.1%) | 23 | 11 (47.8%) | 0.003* |
| LV endocardial infero-basal scar (%) | 33 | 4 (12.1%) | 23 | 9 (39.1%) | 0.019* |
| LV endocardial basal-lateral scar (%) | 33 | 8 (24.2%) | 23 | 12 (52.2%) | 0.032* |
| LV endocardial septal scar (%) | 33 | 5 (15.2%) | 23 | 1 (4.3%) | 0.384 |
| LV endocardial apical scar (%) | 33 | 4 (12.1%) | 23 | 0 (0.0%) | 0.136 |
| RV endocardial scars (%) | 11 | 3 (27.3%) | 4 | 3 (75.0%) | 0.235 |
| RV endocardial free-wall scar (%) | 11 | 1 (9.1%) | 4 | 0 (0.0%) | 1.000 |
| RV endocardial RVOT scar (%) | 11 | 1 (9.1%) | 4 | 2 (50.0%) | 0.154 |
| RV endocardial septal scar (%) | 11 | 2 (18.2%) | 4 | 2 (50.0%) | 0.516 |
| Complications (%) | 41 | 4 (9.8%) | 22 | 1 (4.5%) | 0.650 |

Group 1: patients submitted solely to substrate mapping. Group 2: patients in which functional mapping was performed. was considered. \*P-Value < 0.05. Sample distribution was analyzed by the Kolmogorov-Smirnov test. Categorical variables were compared using the Chi-square test or Fisher's Exact test when appropriate. Numerical variables were compared using Student's t test or Mann-Whitney U test, according to sample distribution. RVOT: Right Ventricle Outflow Tract. VT: Ventricular Tachycardia. LV: Left Ventricle. RV: Right Ventricle.

### Clinical follow-up and outcomes

Considering the complete study population, the total median follow-up time was 6.9 (1.8-20.9) months; when only the outcome “death” is considered for patients who presented VT recurrence and died, median follow-up time reached 8.0 (2.0-21.6) months.

In total, 25 patients (37.9%) experienced VT recurrence during a maximum post-ablation follow-up of 30 months. Among these cases, 5 patients (20.0%) encountered recurrences within the first month, 12 patients (48.0%) within 6 months, and 19 patients (76.0%) experienced VT recurrence within 12 months. The median time to CC-VT recurrence post-ablation was 4.5 months (1.5-10.4). Data on electric storms following ablation were documented for 46 cases (69.7% of the total), with 6 individuals (13.0%) experiencing post-procedure VT recurrence with electric storms.

During the follow-up period, 8 patients died. Among these cases, 5 deaths (62.5%) occurred within the first month following ablation and 7 deaths (87.5%) occurred within 3 months after the procedure. The median time to death after ablation was 0.6 (0.2-1.8) month.

### Predictors of post-ablation VT recurrence

We performed a multivariate analysis to evaluate 30-month post-ablation VT recurrence, with the final model comprising 59 (89.4%) patients, of whom 24 (40.7%) presented ventricular tachycardia recurrence. The multivariate model included the parameters: “age >60 years”, “male gender”, “pre-ablation NYHA class 3 or 4”, “pre-ablation electrical storms”, “pre-ablation incessant VT”, and “performing of electroanatomic functional mapping”. The parameter “pre-ablation LVEF <25%” was used as a strata variable, as discussed in the “Methods” section.

In the final multivariate model, after adjustment to clinically relevant predictors, functional mapping-assisted ablation demonstrated a statistically significant 90.3% reduction in the risk of post-ablation VT recurrence compared to procedures without functional mapping (HR: 0.097, 95% CI: 0.012-0.760, P=0.026). Also, male gender independently associated with decreased risk of VT recurrence during follow-up (HR: 0.354, 95% CI: 0.134-0.940, P=0.037). Importantly, the addition of “Functional mapping” to the final multivariate analysis significantly improved the model (X^2^(1)=9.134, P=0.003). Figure 3 features a forest plot, providing a clearer illustration of the results from the final multivariate analysis.

**Figure 3.**
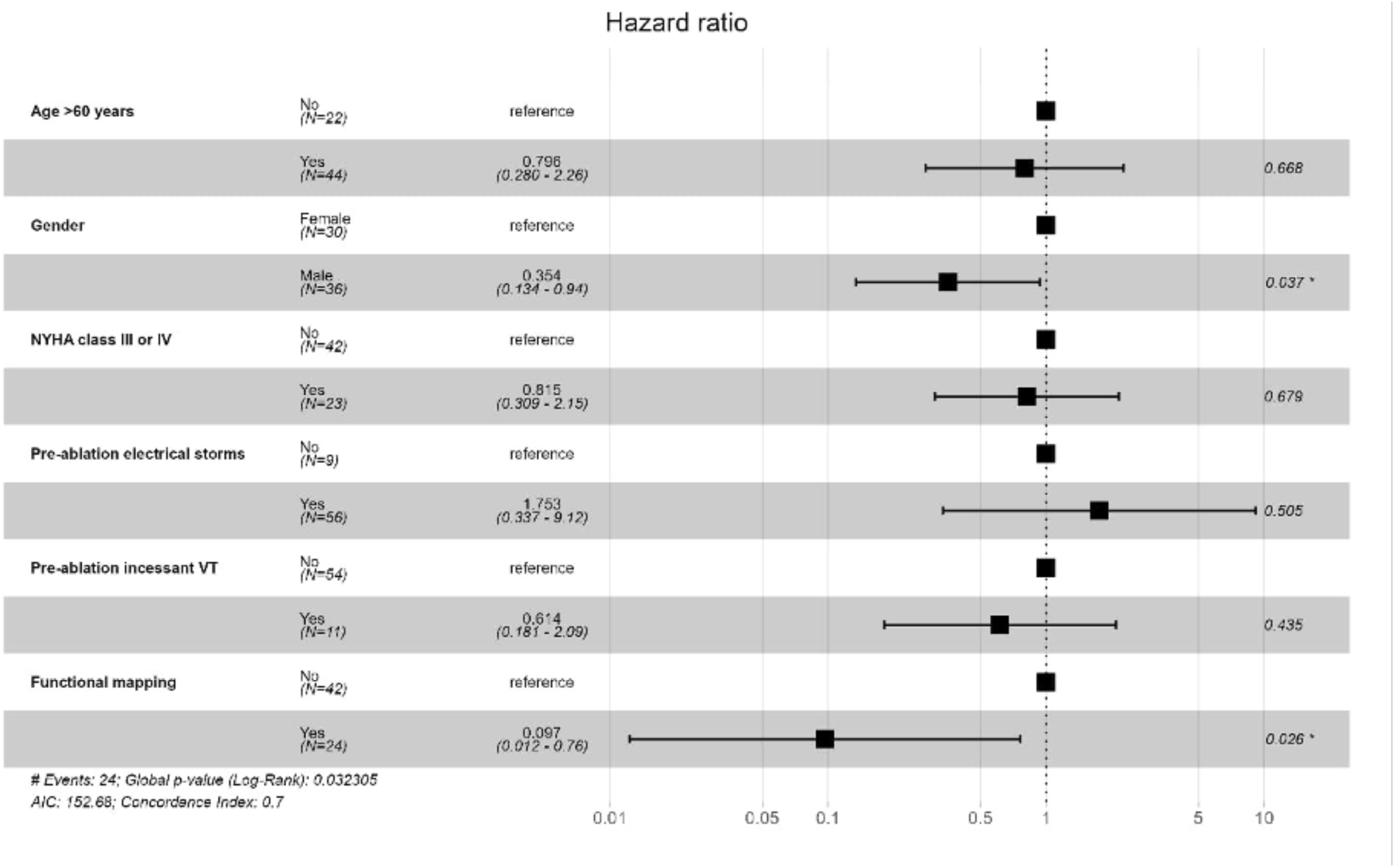
Forest plot illustrating the effect of functional mapping in post-ablation VT recurrence, after adjusting for relevant clinical confounding parameters. NYHA: New York Heart Association; VT: Ventricular Tachycardia; AIC: Akaike Information Criterion.

Finally, Kaplan-Meier analysis revealed a clear and statistically significant reduction in post-ablation VT recurrence when employing functional mapping-assisted procedures (P=0.045). Specifically, when comparing the populations subjected to functional mapping (group 2) and voltage-based mapping (group 1), the VT recurrence-free survival rate at 12 months was 83.2% and 51.9%, respectively. This discrepancy in recurrence-free rates persisted over a 30-month follow-up, with survival rates of 83.2% for functional mapping and 36.4% for voltage-based mapping populations (Figure 4).

**Figure 4.**
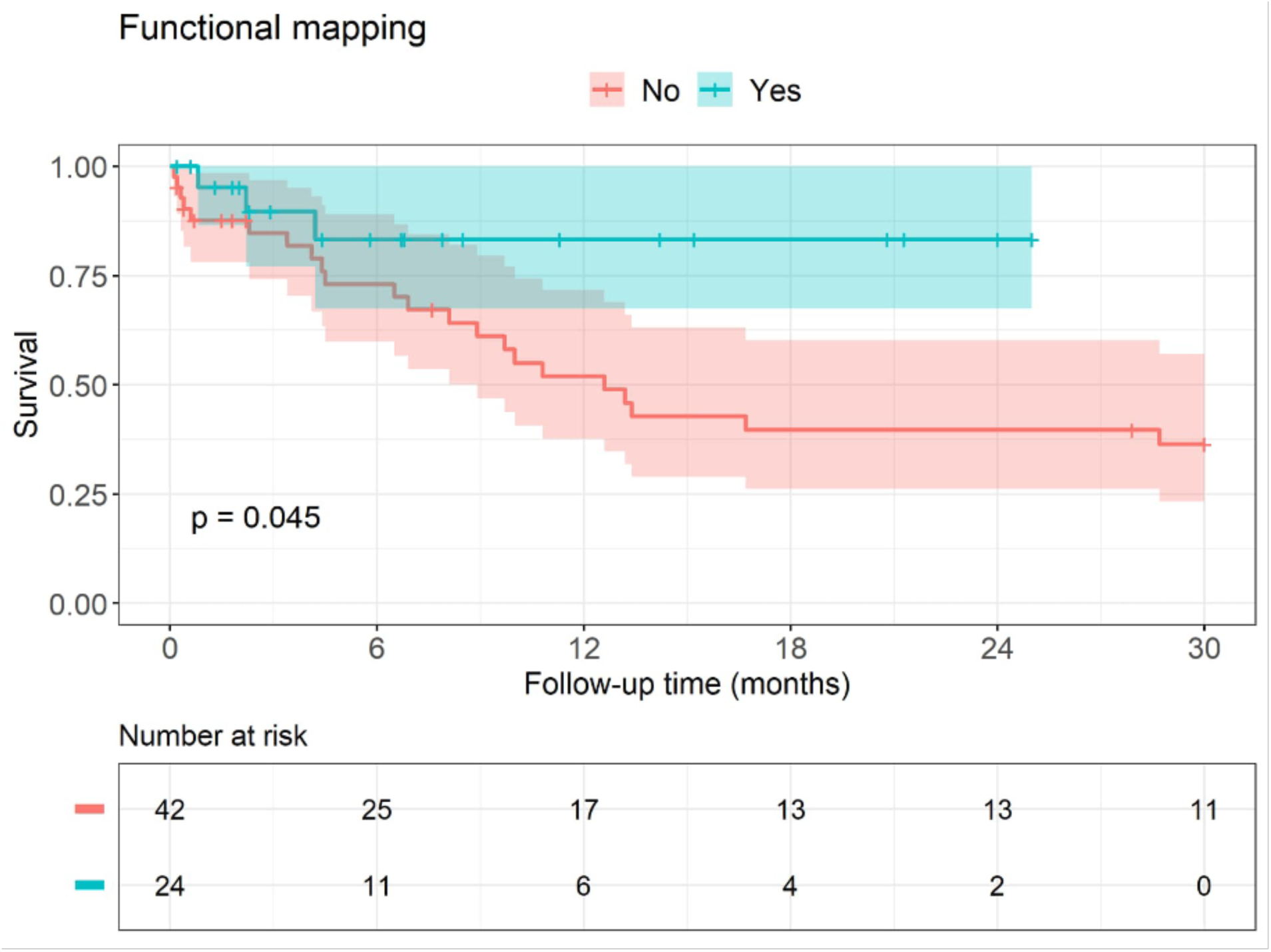
Kaplan-Meier curve evidencing increased recurrence-free survival in the functional mapping group versus voltage-based mapping population. The log-rank test was performed to compare survival between groups.

## Discussion

To the best of our knowledge, this is the first study showing the association of a functional mapping-based strategy to increased rates of survival free of VT recurrence following catheter ablation in a population with highly arrhythmogenic cardiomyopathy. Previous studies have consistently indicated that Chagas cardiomyopathy is associated with elevated rates of VT recurrence and appropriate ICD therapies when compared to other etiologies of cardiomyopathies^2–4^. Given the extensive epicardial and endocardial substrate involved in Chagas VT ablation, it is anticipated that this procedure may carry higher rates of VT recurrence, as indicated in some prior reports^33^.

Our primary finding, which demonstrates reduced rates of VT recurrence in a severe subset of patients, substantiates earlier hypotheses suggesting improved outcomes for patients undergoing functional map-based ablation^34^. It is essential to note that our study cohort comprises patients with highly severe cardiomyopathy, with 86.2% of them experiencing electrical storms prior to ablation. Consequently, strategies aimed at enhancing the efficacy of substrate modification and preventing hemodynamic deterioration are particularly well-suited to improve the overall clinical outcomes in this challenging patient population.

It is important to highlight that, as illustrated in Figure 1, we have developed our own workflow, which incorporates the principles of various functional mapping techniques^5–9^. This workflow was meticulously designed to facilitate a thorough substrate modification while minimizing procedural duration, as longer procedures have been associated with increased mortality^35^. Additionally, we employ unipolar voltage mapping to identify concealed substrates in areas where hidden scar is anticipated, such as the septal and periaortic regions.

While no study to date has demonstrated the superiority of substrate modification over traditional mapping techniques during VT^36,37^, our study directly compares two distinct substrate modification strategies. The first strategy involves working with a fixed anatomical substrate, while the second employs extra-stimulation to uncover concealed regions with decremental properties, functional block, or slow conduction. By initially targeting these areas, our approach aims to achieve a more comprehensive substrate modification through relatively more targeted ablation.

In conjunction with our functional substrate mapping approach for VT ablation, we employed dedicated multipolar catheters to enhance the characterization of local electrograms. This combination enabled a more precise interpretation of near-field signals. While we cannot independently assess the contribution of each component of our approach, the integration of our substrate mapping strategy with multipolar catheters and high-density mapping correlated with a reduced incidence of VT recurrence.

Lastly, our comprehensive mapping strategy enabled us to identify an incidence of 26.8% for the periaortic substrate in patients with Chagas cardiomyopathy referred for VT ablation. This particular substrate localization has not been previously documented in patients with CC submitted to VT ablation. It is worth noting that this substrate is associated with a higher risk of ablation failure^26^. Therefore, comprehensive mapping and ablation of this substrate holds the potential to improve outcomes in this patient population.

Our study presents some limitations. Firstly, there’s a difference in the timing of the ablation procedure between the two analyzed groups. Patients included in Group 1, subjected solely to voltage-based substrate mapping, underwent VT ablation in an earlier time frame compared to Group 2, in which functional mapping was performed. Another limitation is the exclusion of some patients due to a lack of follow-up data. However, once both included and excluded groups showed no statistically significant differences in the major analyzed parameters (Supplementary Table S1), such an issue represents a minor concern. Lastly, the observational nature of this study inherently restricts its capacity to establish causation and susceptibility to various biases that should not be overlooked.

This study also presents several noteworthy strengths. All procedures in both groups were performed collaboratively by two experienced cardiac electrophysiologists, minimizing inter-procedural variability, and ensuring data reproducibility. Moreover, all patients included in the study had VT attributable to Chagas Cardiomyopathy, ensuring a homogeneous population for analysis. Since Chagas Cardiomyopathy is a highly arrhythmogenic condition, and, as demonstrated in our cohort, patients presented with a notably severe baseline clinical profile, it is possible that our analysis may be underestimating the genuine benefits of the described functional mapping strategy for post VT ablation outcomes. Finally, the employment of a robust statistical methodology enabled the formulation of robust hypotheses regarding the benefit of functional mapping techniques for outcomes following VT ablation.

## Conclusion

In summary, our study supports the hypothesis that employing functional mapping strategies in ventricular tachycardia (VT) ablation for Chagas cardiomyopathy may independently reduce the risk of post-ablation VT recurrence in a highly arrhythmogenic population. This finding holds crucial implications given the persistent public health challenge posed by Chagas cardiomyopathy across Latin America, and its recent dissemination across the United States and Europe.

Despite limitations in our study, it is important to stress that such positive results manifested in a population characterized by significant clinical severity and challenging arrhythmogenic substrate, attributes that qualify it as an interesting proof-of-concept for guiding clinical decisions in other arrhythmogenic cardiomyopathies undergoing VT ablation. Furthermore, our findings advocate for the development of double-blind randomized controlled clinical trials to ascertain the actual benefits of functional electroanatomic mapping over substrate mapping in preventing post-ablation ventricular tachycardia recurrence.

## Data Availability

Availability of data and material: The datasets generated and analyzed during the current study are not publicly available due to privacy and ethical considerations. However, de-identified data may be made available from the corresponding author upon reasonable request and approval from the institutional review board. Code availability: The specific algorithms, codes, and software used in data analysis and interpretation are available from the corresponding author upon reasonable request. The authors are committed to transparency and reproducibility, providing necessary information for verification and further research.

## Authors’ contributions and acknowledgments

All authors contributed significantly to the conception, design, analysis, and interpretation of data. B.W., M.C.P.N., and A.A.L.C. conceptualized and designed the analysis. B.C.B., G.A.S., M.P.M., R.C., and A.A.L.C. were involved in data acquisition. B.W., B.C.B., J.L.P.S., M.C.P.N., and A.A.L.C. performed the statistical analysis. B.W., B.C.B., G.A.S., M.P.M., M.P.T.B., A.T.F., M.R.Q.F., C.R.S.A, R.C.M., A.L.P.R., M.C.P.N., and A.A.L.C. were responsible for analysis and data interpretation. The manuscript was drafted and revised by all authors. All authors read and approved the final version of the manuscript. The order of authorship reflects a collective decision regarding the substantial intellectual contribution of each author to the study.

## Supplemental Material

Table S1

Figure S1–S2

