## Supplementary Material for "ENHANCING VENTRICULAR TACHYCARDIA ABLATION OUTCOMES: THE IMPACT OF FUNCTIONAL MAPPING IN CHAGAS CARDIOMYOPATHY"

**SUPPLEMENTAL MATERIAL:**

**SUPPLEMENTARY FIGURE S1-S2:**

Investigation of non-linear effect of numerical variables included in the multivariate model was performed using log hazard splines-adjusted graphical analysis. The dependent variable considered is the study’s end-point: 30-month post-ablation ventricular tachycardia (VT) recurrence rate.

**Supplementary Figure S1.** Pre-ablation left ventricle ejection fraction (LVEF) log hazard splines-adjusted graph for 30-month VT recurrence.
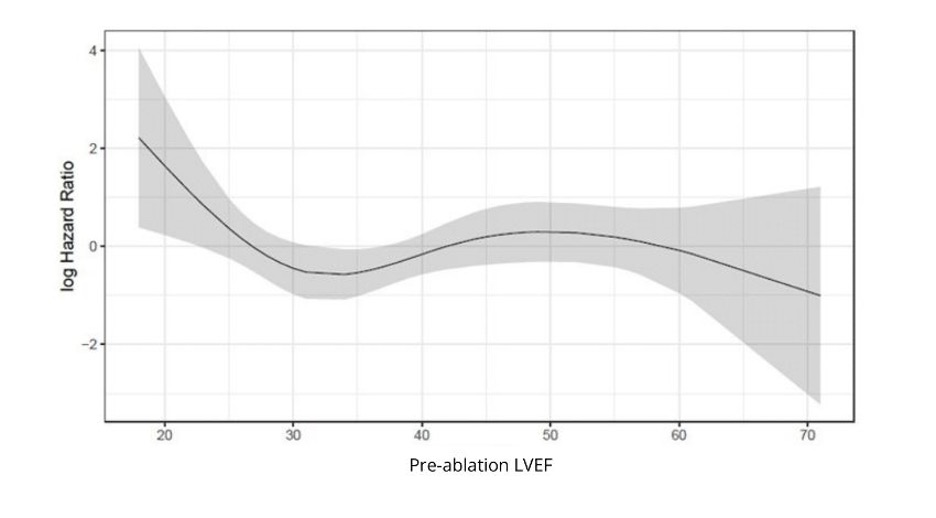

LVEF: Left ventricle ejection fraction.

**Supplementary Figure S2.** Age log hazard splines-adjusted graph for 30-month VT recurrence.

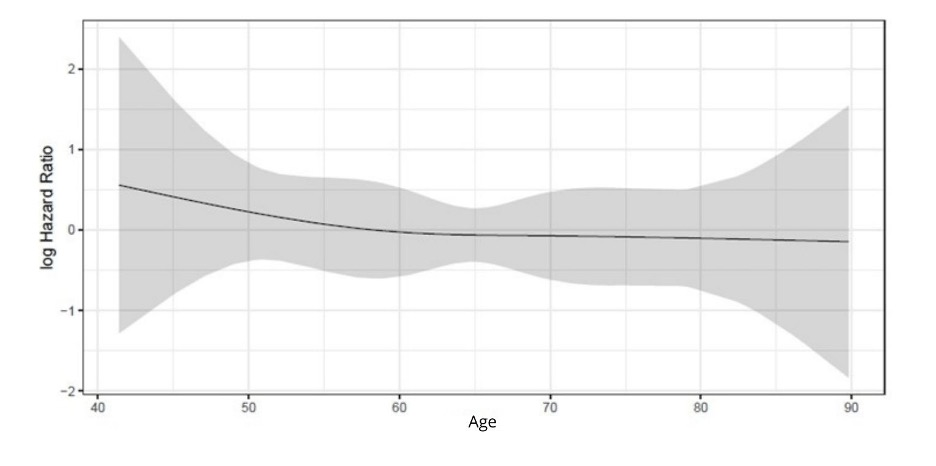

**Supplementary Table S1.** Comparison of baseline characteristics between patients included and excluded of the study.

|  | **Parameters** | | | | **Included (n= 66)** | | | **Excluded (n = 18)** | | ***P*-Value** |
| --- | --- | --- | --- | --- | --- | --- | --- | --- | --- | --- |
|  |  |  |  |  | **N** | **Descriptors** | | **N** | **Descriptors** |  |
| Male gender (%) | | | | | 66 | 36 (54.5%) | | 18 | 12 (66.7%) | 0.428 |
| Implanted device (%) | | | | | 63 | 60 (95.2%) | | 17 | 15 (88.2%) | 0.286 |
| Pre-ablation PAINESD score, median (P25-P75) | | | | | 32 | 8 (5-11) | | 7 | 6 (5-11) | 0.484 |
| Chronic obstructive pulmonary disease (%) | | | | | 32 | 1 (3.1%) | | 7 | 1 (14.3%) | 0.331 |
| Age, median (P25-P75), years | | | | | 66 | 64.2 (57.1-72.8) | | 18 | 64.2 (57.4-71.0) | 0.785 |
|  | Age >60 years (%) | | | | 42 | 44 (66.7%) | | 18 | 12 (66.7%) | 1.000 |
| Ischemic cardiomyopathy (%) | | | | | 32 | 1 (3.1%) | | 7 | 0 (0%) | 1.000 |
| Pre-ablation NYHA class | | | | |  |  | |  |  |  |
|  | NYHA 1 (%) | | | | 65 | 10 (15.4%) | | 18 | 0 (0%) | 0.300 |
|  | NYHA 2 (%) | | | |  | 32 (49.2%) | |  | 9 (50.0%) |  |
|  | NYHA 3 (%) | | | |  | 14 (21.5%) | |  | 6 (33.3%) |  |
|  | NYHA 4 (%) | | | |  | 9 (13.8%) | |  | 3 (16.7%) |  |
| Pre-ablation LVEF, median (P25-P75), % | | | | | 66 | 37.5 (29.8-47.0) | | 18 | 33.5 (27.5-41.0) | 1.161 |
|  | Pre-ablation LVEF <30% (%) | | | | 42 | 16 (24.2%) | | 18 | 4 (22.2%) | 1.000 |
|  | Pre-ablation LVEF <25% (%) | | | | 42 | 7 (10.6%) | | 18 | 4 (22.2%) | 0.238 |
| Pre-ablation electrical storms (%) | | | | | 65 | 56 (86.2%) | | 18 | 14 (77.8%) | 0.465 |
| Diabetes mellitus (%) | | | | | 32 | 3 (9.4%) | | 7 | 1 (14.3%) | 0.563 |
| Number of pre-ablation appropriate ICD therapies, median (P25-P75) | | | | | 61 | 54 (22-98) | | 15 | 45 (13-122) | 0.845 |
| Pre-ablation incessant VT (%) | | | | | 65 | 11 (16.9%) | | 18 | 2 (11.1%) | 0.724 |
| Pre-ablation antiarrhythmic drugs | | | | |  |  | |  |  |  |
|  | Amiodarone (%) | | | | 66 | 61 (92.4%) | | 18 | 13 (72.2%) | 0.033* |
|  | β-blockers (%) | | | | 66 | 64 (97.0%) | | 18 | 17 (94.4%) | 0.520 |
|  | Propafenone (%) | | | | 66 | 1 (1.5%) | | 18 | 0 (0%) | 1.000 |
| Ablation parameters | | | | |  |  | |  |  |  |
|  | Circulatory assistance (%) | | | | 63 | 5 (7.9%) | | 13 | 1 (7.7%) | 1.000 |
|  | Epicardial scars (%) | | | | 57 | 52 (91.2%) | | 12 | 11 (91.7%) | 1.000 |
|  | LV endocardial scars (%) | | | | 56 | 32 (57.1%) | | 12 | 9 (75.0%) | 0.338 |
|  |  | LV endocardial periaortic scar (%) | | | 56 | 15 (26.8%) | | 12 | 2 (16.7%) | 0.716 |
|  |  | LV endocardial infero-basal scar (%) | | | 56 | 13 (23.2%) | | 12 | 6 (50.0%) | 0.080 |
|  |  | LV endocardial basal-lateral scar (%) | | | 56 | 20 (35.7%) | | 12 | 7 (58.3%) | 0.197 |
|  |  | LV endocardial septal scar (%) | | | 56 | 6 (10.7%) |  | 12 | 1 (8.3%) | 1.000 |
|  |  | LV endocardial apical scar (%) | | | 56 | 4 (7.1%) |  | 12 | 3 (25.0%) | 0.099 |
|  | RV endocardial scars (%) | |  | | 15 | 6 (40.0%) |  | 0 | 0 (0%) | - |
|  |  | RV endocardial free-wall scar (%) | | | 15 | 1 (6.7%) |  | 0 | 0 (0%) | - |
|  |  | RV endocardial RVOT scar (%) | | | 15 | 3 (20.0%) |  | 0 | 0 (0%) | - |
|  |  | RV endocardial septal scar (%) | | | 15 | 4 (26.7%) |  | 0 | 0 (0%) | - |
|  | Complications (%) | | |  | 63 | 5 (7.9%) |  | 13 | 1 (7.7%) | 1.000 |

*P-Value <0.05. Sample distribution was analyzed by the Kolmogorov-Smirnov test. Categorical variables were compared using the Chi-square test or Fisher’s Exact test when appropriate. Numerical variables were compared using Student’s t test or Mann-Whitney U test, according to sample distribution. LVEF: Left Ventricle Ejection Fraction. NYHA: New York Heart Association functional class. VT: Ventricular Tachycardia. LV: Left Ventricle. RV: Right Ventricle. RVOT: Right Ventricle Outflow Tract. P25-P75: Percentile 25 - Percentile 75. PAINESD Score: chronic cardiomyopathy, NYHA class 3 or 4, LVEF <25%, VT storm, and diabetes mellitus.
